# Comparison of endoscopic activity before and during the covid pandemic at a tertiary care hospital in South Punjab

**DOI:** 10.1101/2021.10.11.21264820

**Authors:** Farooq Mohyud Din Chaudhary, Muhammad Asif Gul, Nouman Hameed, Rizwan Hameed, Yasir Zaidi, Shehryar Kanju, Ahsan Tameez-ud-din, Syeda Manal Altaf, Asma Tameez Ud Din

**Affiliations:** Department of Gastroenterology, Nishtar Medical University & Hospital, Multan, Pakistan; Department of Dermatology, Benazir Bhutto Hospital, Rawalpindi, Pakistan; Rawalpindi Medical University, Rawalpindi, Pakistan; Department of Hematology & Pathology, Shahida Islam Medical College, Lodharan, Pakistan

**Author notes:** Corresponding Author: Farooq Mohyud Din Chaudhary, Email Address, Contact number, Address: House no. 129, Street no. 8, Pearl City, Multan Cantt, Multan. **Funding:** No funding was received for the publication of this article.

**Keywords:** MeSH, Endoscopy, COVID-19, Gastroenterology

## Abstract

**Introduction:** Coronavirus disease 2019 (COVID-19) has resulted in dramatic changes to health-care delivery. Endoscopic activity has had frequent disruptions during this pandemic. The objective of the study was to see the influence of pandemic over the endoscopic activity.

**Methods:** This retrospective analysis of endoscopic activity was undertaken at Nishtar Hospital Multan. Procedural analysis was done in the three months immediately after covid lockdown (1st April till 30th June 2020) and was compared to a similar period one year back.

**Results:** Five hundred and fifty-four (68.5%) patients underwent endoscopic procedures during the three months of pre-COVID era, while this number reduced to half (n=255, 31.5%) patients during the covid pandemic. Even though the absolute number of Esophagogastroduodenoscopies (EGDs) reduced during the pandemic, patients were more likely to undergo EGDs during the COVID pandemic in contrast to the era before the pandemic (79% versus 66%, p = 0.002). The most common indication for EGD was upper gastrointestinal bleeding (UGIB). The percentage of EGDs done for UGIB rose from almost 60% to 80% during the covid pandemic (p < 0.001). The most common findings were esophageal varices and portal gastropathy (non-significant difference during and before the pandemic). Percentage of ERCPs done for obstructive jaundice doubled during the COVID pandemic (33% versus 65%, p = 0.002).The most common indication for sigmoidoscopy or colonoscopy was lower gastrointestinal bleeding. However, no significant difference was found before and during the covid pandemic (41.7% and 45.8% respectively, p=0.72). Internal hemorrhoids were the most common endoscopic finding. Colon cancer diagnosis reduced from 10% to undetected during the pandemic period.

**Conclusion:** COVID pandemic resulted in considerable reduction in all type of endoscopic procedures. Majority of procedures were done for emergency indications like gastrointestinal bleeding. Rates of cancer detection was significantly reduced.

## INTRODUCTION

Coronavirus disease 2019 (COVID-19) is caused by severe acute respiratory syndrome coronavirus-2 (SARS-CoV-2). Within a short span of time, the disastrous impact of this pandemic has reached unprecedented levels, affecting countries all over the world with more than 206 million reported cases and more than 4.3 million deaths as of 8th August 2021 [1]. COVID-19 has greatly impacted medical practices of health-care centers including private clinics and tertiary care hospitals. The field of Gastroenterology has been especially affected and to meet the challenges posed by this pandemic, guidelines on safe endoscopy have been published by esteemed United States, European, Asian and Japanese societies [2-5].

It is a well-recognized fact that COVID-19 is primarily transmitted via the respiratory route. However, endoscopists face the risk of transmission via the oral and fecal routes as well [6-9].All over the world gastrointestinal (GI) departments were affected severely. In most centers elective procedures were postponed which led to an unprecedented decrease in the endoscopic activity [10-13]. The first known case of COVID-19 in Pakistan was reported on 26th February 2020, with lockdown imposed by the Government of Pakistan in March 2020 [14]. The provision of essential and non-essential health services in the country have been severely affected by the pandemic but few studies regarding the impact of this pandemic over the endoscopic activity have been reported in Pakistan. We aimed to investigate how the COVID pandemic has influenced the endoscopic procedures at Nishtar Hospital Multan, which is the largest tertiary care center in the region.

## METHODS

### Scope of study

This retrospective study was carried out at the Department of Gastroenterology, Nishtar Hospital Multan. Ethical approval for the study was issued by the Institutional Review Board (IRB) of Nishtar Medical University, Multan (Reference No. 17997). To see the effects of pandemic, a comparison between two time periods was considered, namely 1st of April-30th June 2020 during the pandemic, and a similar period between 1st of April-30th June 2019 before the pandemic.

### Data Collection

All the patients who underwent endoscopic procedures during this time period were included in our study. From the endoscopy record register patient’s age, gender, type of procedure, indication of procedure and findings of endoscopy were noted. Confidentiality of patients was ensured.

### Data Analysis

Data were entered and evaluated in Statistical Package for Social Sciences (SPSS) version 20 (IBM Corp, Armonk, US). Comparison of endoscopic activity was done between the two time periods. The results were reported as frequencies, percentages, and tables. Chi-square test was used for the analysis of qualitative variables. We considered a p-value of less than 0.05 to be significant.

## RESULTS

A total of 809 (males 56.7%, females 43.3%) patients were included in the study who underwent endoscopic procedures during the specified time periods. There were 554 (68.5%) patients who underwent procedures during the three months of pre-COVID era. During the COVID era this number became less than half i.e., only 255 (31.5%) patients. The mean age of the study population was 46.86 years with a standard deviation of 16.6 years and age range of 12 to 90 years. The mean age of patients undergoing procedures during the pre-COVID era was 46.07 ± 16.4 years, while mean age of patients during the COVID-19 pandemic was 48.57 ± 16.9 years (p < 0.05). There was no significant difference with regards to gender distribution between the pre-COVID and COVID pandemic (p = 0.76).

Table 1 shows the different procedures and their frequency before and during the COVID pandemic. Overall EGDs were the most common procedure. Patients were more likely to undergo EGDs during the COVID pandemic as compared to the era before the pandemic (79% versus 66%, p = 0.002).

**Table 1.** Impact of COVID-19 on the number of Endoscopic procedures.

| Type | Pre COVID-19 Time<br>554 | COVID-19 Time<br>255 | P value |
| --- | --- | --- | --- |
| SFS, n (%) | 46 (8.3) | 11 (4.3) | 0.04 |
| CFS, n (%) | 57 (10.3) | 13 (5.1) | 0.01 |
| ERCP, n (%) | 84 (15.2) | 29 (11.4) | 0.15 |
| EGDs, n (%) | 367 (66.2) | 202 (79.2) | <0.001 |

EGD: Esophagogastroduodenoscopy; ERCP: Endoscopic retrograde cholangiopancreatography;
SFS: Sigmoidoscopy; CFS: Colonoscopy.

Table 2 shows the indications and findings of EGDs before and during the covid pandemic. The most common indication for EGD was upper gastrointestinal bleeding (UGIB). The percentage of EGDs done for UGIB rose from almost 60% to 80% during the covid pandemic (p < 0.001). Percentage of EGDs done for persistent pain epigastrium decreased significantly to half (10% to 5%, p < 0.001) during the pandemic. Similarly, EGDs done for variceal screening reduced significantly from 10% to 1% during the pandemic (p< 0.001). Not much change was seen in other indications for which EGD was performed. As for the findings, the two most common seen in almost of EGDs were esophageal varices and portal gastropathy.

**Table 2.**
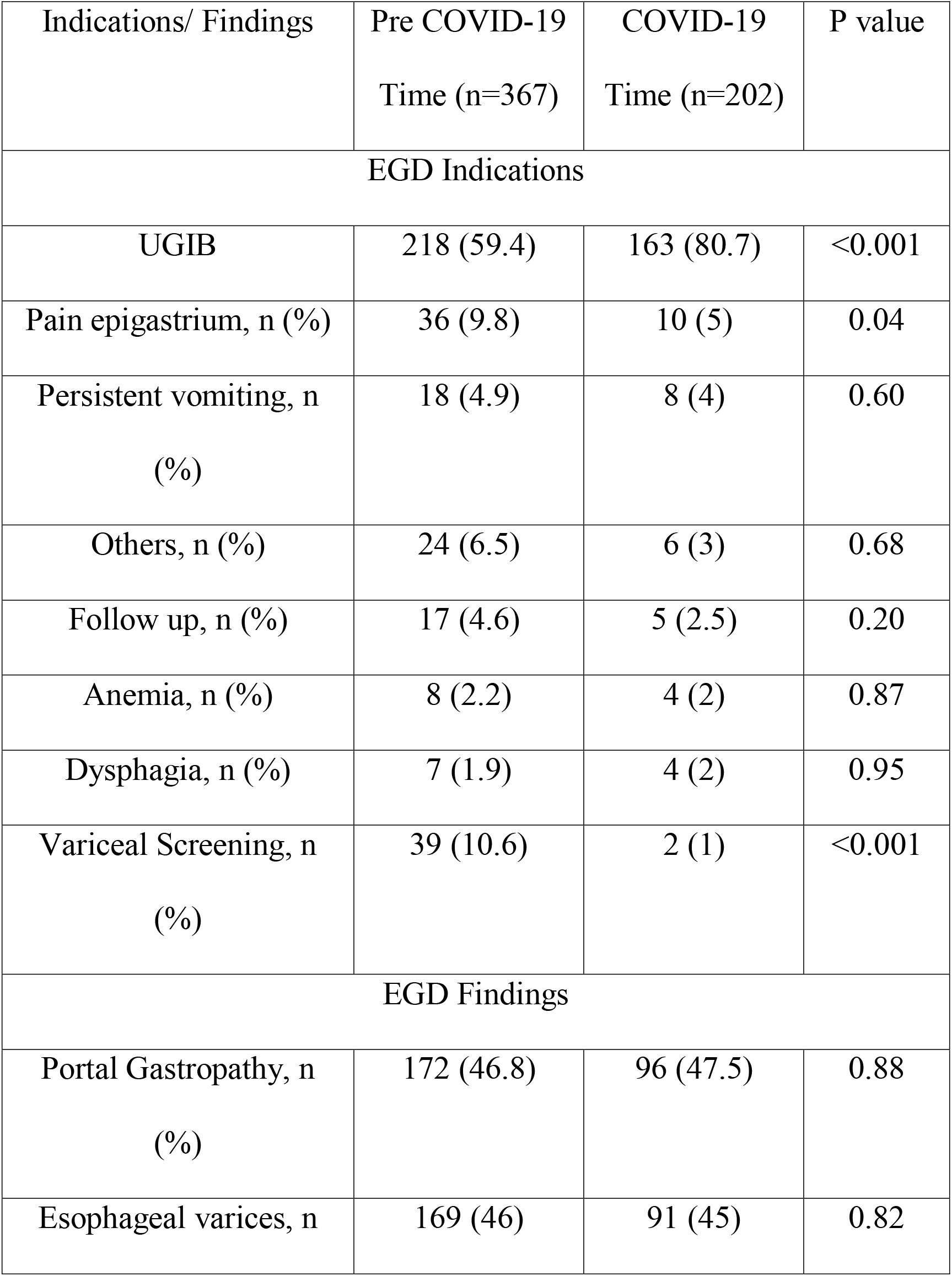
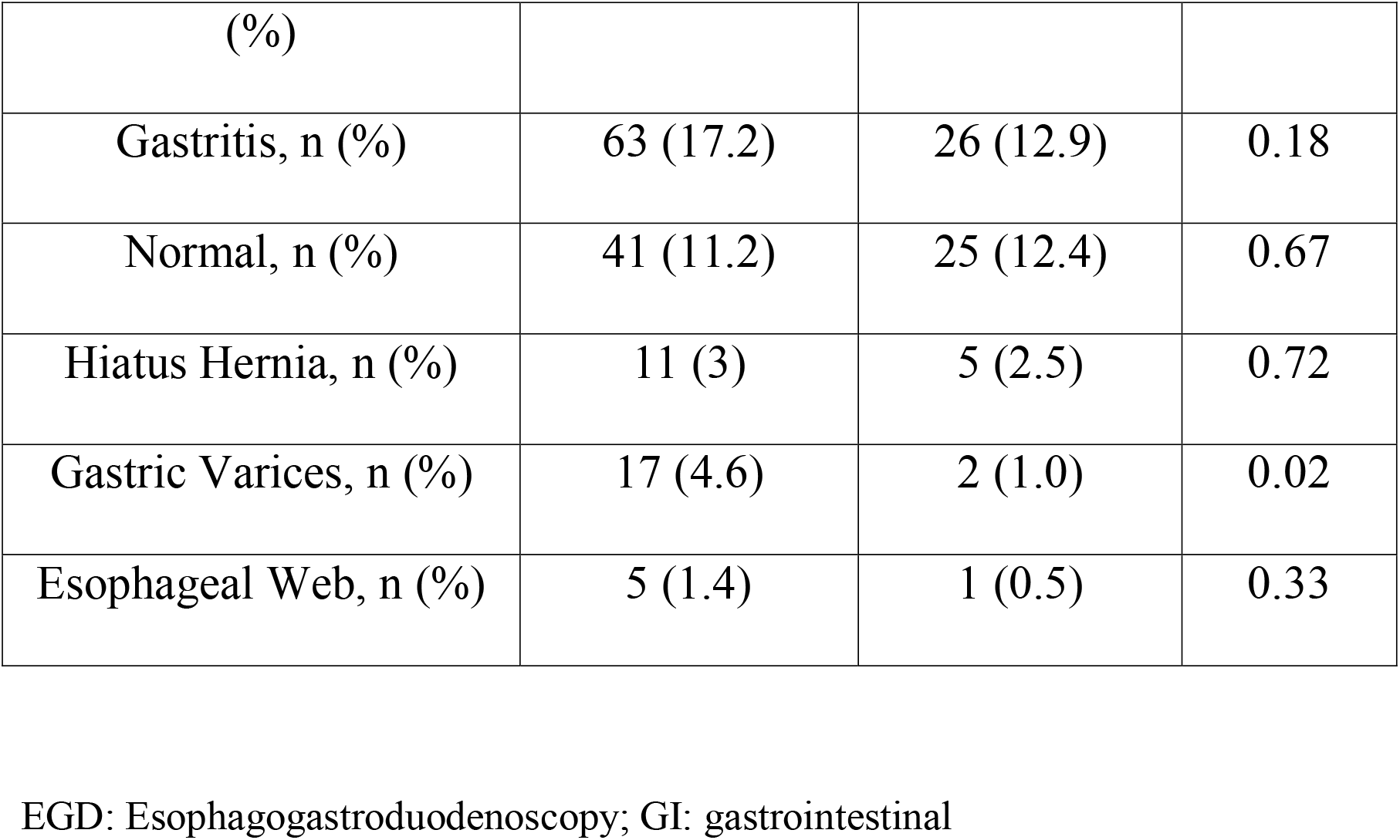
Analysis of EGDs before and during the COVID-19 period.

Table 3 shows the different indications for which ERCP was performed. Obstructive jaundice was the most common indication. Percentage of ERCPs done for obstructive jaundice doubled during the COVID pandemic (33% versus 65%, p = 0.002). Table 4 shows the indications and findings of lower GI endoscopy procedures performed. The most common indication for sigmoidoscopy or colonoscopy was lower gastrointestinal bleeding. However, no significant difference before and during the covid pandemic (41.7% during the pre-COVID era, 45.8% during the COVID pandemic, p = 0.72). Internal hemorrhoids were the most common endoscopic finding in patients who underwent lower GI endoscopy (38% in pre-COVID era, 50% during the pandemic).

**Table 3.** Analysis of ERCPs before and during the COVID-19 period.

| Indications | Pre COVID-19 Time<br>(n=84) | COVID-19 Time (n=29) | P value |
| --- | --- | --- | --- |
| ERCP Indications |  |  |  |
| Obstructive Jaundice | 28 (33.3) | 19 (65.5) | 0.002 |
| Choledocholithiasis | 28 (33.3) | 7 (24.1) | 0.36 |
| CBD Stricture | 10 (11.9) | 2 (6.9) | 0.45 |
| Biliary Pancreatitis | 6 (7.1) | 0 | Not Valid |
| Cholangiocarcinoma | 5 (6) | 0 | Not valid |

ERCP: Endoscopic retrograde cholangiopancreatography; CBD: Common Bile Duct

**Table 4.** Analysis of Lower GI Endoscopies before and during the COVID-19 period.

| Indications/<br>Findings | Pre COVID-19<br>Time (n=103) | COVID-19<br>Time (n=24) | P value |
| --- | --- | --- | --- |
| Lower GI Endoscopy Indications |  |  |  |
| Lower GI<br>Bleeding | 43 (41.7) | 11 (45.8) | 0.72 |
| Altered bowel<br>habits | 20 (19.4) | 1 (4.2) | 0.70 |
| Chronic pain<br>abdomen | 26 (25.2) | 7 (29.1) | 0.69 |
| CRC screening | 4 (3.9) | 4 (16.7) | 0.02 |
| Others* | 10 (9.7) | 1 (4.2) | 0.38 |
| Lower GI Endoscopy Findings |  |  |  |
| Internal<br>Hemorrhoids | 39 (37.9) | 12 (50) | 0.27 |
| Colitis | 24 (23.3) | 5 (20.8) | 0.80 |
| Normal | 24 (23.3) | 6 (25) | 0.86 |
| Cancer | 11 (10.7) | 0 (0) | Not Valid |
| Rectal Polyp | 1 (0.98) | 1 (4.2) | 0.26 |
| Rectal Ulcer | 1 (0.98) | 0 (0) | Not Valid |
| Strictures | 1 (0.98) | 0 (0) | Not Valid |

GI: gastrointestinal; CRC: Colorectal Cancer;
\*Others: (Anemia, painful defecation, mucous discharge, ileostomy reversal)

## DISCUSSION

Coronavirus pandemic has shaken the global health infrastructure. Third world countries with an already burdened health sector (like Pakistan) have felt the full impact of the pandemic as their burgeoning economic woes have made the provision of a comprehensive health care during this period close to impossible. Gastroenterology is one of the worst affected fields of medicine in the country but there is a paucity of literature regarding this topic so we analyzed the impact of covid-19pandemic on the endoscopic activity at a large government hospital in South Punjab in order to make the relevant health authorities aware of the extent of the impact of the pandemic over this field.

The results of our study indicated that there was a significant decrease in overall number of procedures during the covid-19 pandemic. This is similar to what has been reported in studies from other parts of the world [11, 15-17]. A significant reduction in the number of sigmoidoscopies, colonoscopies and OGDs was seen. A non-significant reduction was seen in ERCPs during the covid pandemic. Rutter et al. described similar results from UK. The authors reported marked reduction in sigmoidoscopies, colonoscopies and OGDs, but only 44% reduction in ERCPs [11].

In our setting, it was found that although the overall absolute number of procedures reduced to almost half during the covid pandemic, there were some interesting findings with regards to the indications for these procedures. There was an increase in the percentage of EGDs and the proportion ERCP procedures for obstructive jaundice doubled during this period (p = 0.002). These changes occurred due to precautionary restrictions adopted by the government and our institute, limiting the procedures mostly to emergency indications and cancer screening. Thus, indications such as pain epigastrium, vomiting, follow up, variceal screening and altered bowel habits were considerably decreased. Our results are comparable to a multicenter study from Italy that reported a reduction in total number of endoscopic procedures which varied from no reduction to 100% [13]. The considerable reduction in endoscopic activity resulted in a decrease in the number of cancer detection especially of the colon. This is consistent with the global trend of cancer detection during the covid pandemic [11, 13, 16].

Most endoscopy units across the globe have resumed elective GI procedures. However, the different waves of coronavirus variants threaten the normalization of endoscopic activity as it was before this pandemic. Sustaining elective GI procedures is important otherwise it may have drastic effects on the health of patients by delaying the diagnosis of cancer and other life-threatening conditions [18].

This study is one of the first in this region to evaluate the effect of covid pandemic over endoscopy practice but it has certain limitations. Our study was single centered and the database contained no information regarding complications of different procedures.

## CONCLUSION

The covid pandemic has had a profound impact on the endoscopic activity, which is evident by a considerable reduction in all types of procedures. Most procedures were carried out in emergency settings and cancer detection was reduced during these times. Only time will tell the full impact of covid pandemic and further studies are required to delineate the long-term effect of this health disaster over the well-being of the patients. Efforts should be done to safely return to pre-covid situation so that the patients in need of essential health services can be evaluated and treated in the way which suits their health needs.

## Data Availability

All data produced in the present work are contained in the manuscript.

## REFERENCES

1. Worldometer August 8th. Coronavirus Update (Live). Available online at: https://www.worldometers.info/coronavirus/#countries

2. Chiu PWY, Ng SC, Inoue H, Reddy DN, Ling Hu E, Cho JY, Ho LK, Hewett DG, Chiu HM, Rerknimitr R, Wang HP, Ho SH, Seo DW, Goh KL, Tajiri H, Kitano S, Chan FKL. Practice of endoscopy during COVID-19 pandemic: position statements of the Asian Pacific Society for Digestive Endoscopy (APSDE-COVID statements). Gut 2020; 69: 991–996 [PMID: 32241897 DOI: 10.1136/gutjnl-2020-321185]

3. Gralnek IM, Hassan C, Beilenhoff U, Antonelli G, Ebigbo A, Pellisè M, Arvanitakis M, Bhandari P, Bisschops R, Van Hooft JE, Kaminski MF, Triantafyllou K, Webster G, Pohl H, Dunkley I, Fehrke B, Gazic M, Gjergek T, Maasen S, Waagenes W, de Pater M, Ponchon T, Siersema PD, Messmann H, Dinis-Ribeiro M. ESGE and ESGENA Position Statement on gastrointestinal endoscopy and the COVID-19 pandemic. Endoscopy 2020; 52: 483–490 [PMID: 32303090 DOI: 10.1055/a-1155-6229]

4. Irisawa A, Furuta T, Matsumoto T, Kawai T, Inaba T, Kanno A, Katanuma A, Kawahara Y, Matsuda K, Mizukami K, Otsuka T, Yasuda I, Tanaka S, Fujimoto K, Fukuda S, Iishi H, Igarashi Y, Inui K, Ueki T, Ogata H, Kato M, Shiotani A, Higuchi K, Fujita N, Murakami K, Yamamoto H, Ito T, Okazaki K, Kitagawa Y, Mine T, Tajiri H, Inoue H. Gastrointestinal endoscopy in the era of the acute pandemic of coronavirus disease 2019: Recommendations by Japan Gastroenterological Endoscopy Society (Issued on April 9th, 2020). Dig Endosc 2020; 32: 648–650 [PMID: 32335946 DOI: 10.1111/den.13703]

5. Sultan S, Lim JK, Altayar O, Davitkov P, Feuerstein JD, Siddique SM, Falck-Ytter Y, El-Serag HB; AGA Institute. AGA Rapid Recommendations for Gastrointestinal Procedures During the COVID-19 Pandemic. Gastroenterology 2020; 159: 739–758. e4 [PMID: 32247018 DOI: 10.1053/j.gastro.2020.03.072]

6. Konturek PC, Harsch IA, Neurath MF, Zopf Y. COVID-19 - more than respiratory disease: a gastroenterologist’s perspective. J Physiol Pharmacol 2020; 71 [PMID: 32633236 DOI:10.26402/jpp.2020.2.02]

7. Soetikno R, Teoh AYB, Kaltenbach T, Lau JYW, Asokkumar R, Cabral-Prodigalidad P, Shergill A. Considerations in performing endoscopy during the COVID-19 pandemic. Gastrointest Endosc 2020; 92: 176–183 [PMID: 32229131 DOI: 10.1016/j.gie.2020.03.3758]

8. Zhang J, Wang S, Xue Y. Fecal specimen diagnosis 2019 novel coronavirus-infected pneumonia. JMed Virol 2020; 92: 680–682 [PMID: 32124995 DOI: 10.1002/jmv.25742]

9. Li LY, Wu W, Chen S, Gu JW, Li XL, Song HJ, D. F, Wang G, Zhong CQ, Wang XY, Chen Y,Shah R, Yang HM, Cai Q. Digestive system involvement of novel coronavirus infection: Prevention and control infection from a gastroenterology perspective. J Dig Dis 2020; 21: 199–204 [PMID: 32267098 DOI: 10.1111/1751-2980.12862]

10. Alboraie, M.; Piscoya, A.; Tran, Q.T.; Mendelsohn, R.B.; Butt, A.S.; Lenz, L.; Alavinejad, P.; Emara, M.H.; Samlani, Z.; Altonbary, A.; et al. The global impact of COVID-19 on gastrointestinal endoscopy units: An international survey of endoscopists. Arab. J. Gastroenterol. 2020, 21, 156–161.

11. Rutter, M.D.; Brookes, M.; Lee, T.J.; Rogers, P.; Sharp, L. Impact of the COVID-19 pandemic on UK endoscopic activity and cancer detection: A National Endoscopy Database Analysis. Gut 2020, 1–7.

12. Arantes, V.N.; Martins, B.C.; Seqatto, R.; Milhomen-Cardoso, D.M.; Franzini, T.P.; Zuccaro, A.M.; Alves, J.S.; Maluf-Filho, F. Impact of coronavirus pandemic crisis in endoscopic clinical practice: Results from a national survey in Brazil. Endosc. Int. Open. 2020, 8, E822–E829.

13. Repici, A.; Pace, F.; Gabbiadini, R.; Colombo, M.; Hassan, C.; Dinelli, M.; ITALIAN GI-COVID19 Working Group. Endoscopy Units and the Coronavirus Disease 2019 Outbreak: A Multicenter Experience from Italy. Gastroenterology 2020, 159, 363–366.

14. Government of Pakistan. Coronavirus in Pakistan. http://covid.gov.pk/. Accessed August 16, 2020.

15. Forbes, N.; Smith, Z.L.; Spitzer, R.L.; Keswani, R.N.; Wani, S.B.; Elmunzer, B.J.; North American Alliance for the Study of Digestive Manifestations of COVID-19. Changes in Gastroenterology and Endoscopy Practices in Response to the Coronavirus Disease2019 Pandemic: Results from a North American Survey. Gastroenterology 2020, 159, 772–774.

16. Dinmohamed, A.G.; Visser, O.; Verhoeven, R.H.A.; Louwman, M.W.J.; van Nederveen, F.H.; Willems, S.M.; Merkx, M.A.W.; Lemmens, V.E.P.P.; Nagtegaal, I.D.; Siesling, S. Fewer cancer diagnoses during the COVID-19 epidemic in the Netherlands. Lancet Oncol. 2020, 21, 750–751.

17. Zhu, L.; Cai, M.Y.; Shi, Q.; Wang, P.; Li, Q.L.; Zhong, Y.S.; Yao, L.Q.; Zhou, P.H. Analysis of selective endoscopy results during theepidemic of coronavirus disease 2019 (COVID-19). Zhonghua Wei Chang. Wai Ke Za Zhi. 2020, 23, 327–331.

18. Kushnir, V.M.; Berzin, T.M.; Elmunzer, B.J.; Mendelsohn, R.B.; Patel, V.; Pawa, S.; Smith, Z.L.; Keswani, R.N.; North American Alliance for the Study of Digestive Manifestations of COVID-19. Plans to Reactivate Gastroenterology Practices Following the COVID-19 Pandemic: A Survey of North American Centers. Clin. Gastroenterol. Hepatol. 2020, 18, 2287–2294.

